# Factors influencing focused practice: A qualitative study of resident and early-career family physician practice choices

**DOI:** 10.1101/2021.06.24.21259486

**Authors:** Monisha Kabir, Ellen Randall, Goldis Mitra, M. Ruth Lavergne, Ian Scott, David Snadden, Lori Jones, Laurie J. Goldsmith, Emily G. Marshall, Agnes Grudniewicz

## Abstract

**Background:** Although focused practice within family medicine may be increasing globally, there is limited research on the factors contributing to decisions to focus practice. We aimed to examine the factors influencing resident and early-career family physician choices of focused practice across three Canadian provinces.

**Methods:** We analyzed a subset of qualitative interview data from a study across British Columbia, Ontario, and Nova Scotia. A total of 22 resident family physicians and 38 early-career family physicians in their first 10 years of practice who intend to or currently practice in a focused area were included in our analysis. We compared participant types, provinces, and the degree of focused practice while identifying themes related to factors influencing the pursuit of focused practice.

**Results:** We identified three key themes of factors contributing to choices of focused practice: self-preservation within the current health care system, support from colleagues, and experiences in medical school and/or residency. Minor themes included alignment of practice with skills, personal values, or ability to derive professional satisfaction; personal lived experiences; and having many attractive opportunities for focused practice.

**Interpretation:** Both groups of participants unanimously viewed focused practice as a way to circumvent the burnout or exhaustion they associated with comprehensive practice in the current structure of the health care. This finding, in addition to other influential factors, was consistent across the three provinces. More research is needed to understand the implications of resident and early-career family physician choices of focused practice within the physician workforce.

## Introduction

In recent decades, there has been a global decline in offering a comprehensive scope of practice in family medicine,[1-6] and a concurrent trend towards focused practice,[7-12] wherein one or more specific clinical areas form a major part-time or full-time component of practice.[13] Previous research suggests that this trend is in part due to perceptions that focused practice offers a desirable intellectual challenge[8] and better remuneration.[14] Other studies suggest that the exodus from comprehensive family medicine practice can be attributed to both the breadth and the overwhelming nature of its scope,[7, 12] and undesirable post-training working environments.[3] These studies have not provided an in-depth examination of the factors influencing the pursuit of focused practice in family medicine.

The objective of this study was to examine the factors contributing to choices of focused practice in three Canadian provinces. In Canada, medical school is graduate entry, lasts three to four years, and for family practitioners, is followed by two years as a resident family physician (FP) before certification as an independent family practitioner. We present findings from both resident FPs and independent family practitioners in their first decade of practice, henceforth “early-career FPs”, to address our research question.

## Methods

### Study design and population

We report on a subset of the qualitative interview data collected through a larger mixed methods study examining factors contributing to practice intentions and choices among resident and early-career FPs across British Columbia, Ontario, and Nova Scotia.[15]

We recruited resident and early-career FPs for the qualitative arm of this larger mixed methods study through family medicine residency programs, social media, research conferences, and selected provincial medical associations.

Interested participants completed an online screening survey that captured demographic information and practice characteristics (see Appendix 1). We used purposeful sampling to maximize variation across self-identified gender, marital status, dependents, training location, years of training, years in practice, scope of practice, and practice models in each province. Selected individuals were then invited to participate in a 60-minute interview. We provided participants with study information and an honorarium.

We interviewed 31 of 32 resident FPs and 63 of 69 early-career FPs who had been invited to participate in the larger mixed methods study. Reasons for nonparticipation included scheduling conflicts (n=2), no response (n=4), or withdrawal with no reason provided (n=1). The study received ethics approval in all three provinces.

### Context for this paper

The sample used in this paper consists of 22 resident FPs and 38 early-career FPs who: i) self-identified in the screening survey as intending to practice or currently practicing within a focused area, and/or ii) described focused practice elements in their overall practice during the interview. We considered resident and early-career FPs to have a focused practice, in whole or in part, if a component of their intended or current practice was narrowed or specialized in scope (e.g., addictions medicine, emergency medicine) and they did not intend to or currently deliver any routine comprehensive family medicine care to patients in that part of their practice. We considered resident and early-career FPs to be engaging in comprehensive family medicine practice when they described providing clinic-based, first-contact, longitudinal, comprehensive, and coordinated services to a defined group of patients to address the majority of their health care needs.[16, 17] Our definition of comprehensive family medicine practice included providing comprehensive care for a particular population (e.g., refugees) or comprehensive care omitting obstetrics/prenatal care.

### Data collection

One research analyst (ER, LJ, MM) per province conducted one-on-one, semi-structured, in-depth interviews. Each research analyst was trained in qualitative interviewing. Telephone interviews were conducted using a semi-structured interview guide specific to each subgroup (see Appendix 2). Interviews were audio-recorded and transcribed verbatim. Research analysts recorded their reflections and interview summaries after each interview. Participant recruitment occurred iteratively until no new themes were identified in interviews.

### Data analysis

We used iterative, inductive thematic analysis.[18] For the qualitative arm of the larger mixed methods study, three research analysts (ER, LJ, MM) with experience in qualitative analysis generated initial resident and early-career FP codebooks through inductive coding of one resident and one early-career FP interview.[19] Codebooks were then refined through application to a subset of transcripts with guidance from the senior author (AG). The research analysts used the final codebooks to code transcripts from their respective provinces in NVivo 12[20]. Codebooks were iteratively amended to incorporate emerging codes and ensure consistency between the resident and early-career FP codebooks. The research analysts also coded one interview from another province to ensure reliability.

Data analysis for this paper involved the first author (MK) reviewing interview excerpts coded as influential factors for practice choices and aggregating them into potential overarching themes, based on the identification of patterns between and across the transcripts. We conducted comparative analysis[21] to compare thematic patterns identified from the early-career FP transcripts with the resident FP dataset. If new themes were identified from the resident FP transcripts, we used an iterative approach to find corresponding themes in the early-career FP transcripts. We also compared themes across provinces.

We used accepted strategies for validating qualitative research.[22] The findings were: i) triangulated across a large sample size of participants with diverse experiences from three provinces and two types of participant groups;[23] and ii) presented to FP members of the research team (DS, IS, GM), who were not involved in the analysis process for this paper, to confirm data interpretation.

## Results

### Description of interview participants’ focused practice intentions or choices

The 22 resident and 38 early-career FPs reported a variety of intended or current clinical areas of practice, with most participants reporting a combination of focused area(s) and/or some form of comprehensive practice and focused practice. We present demographic and practice characteristics (Table 1 and 2) to reflect the range of resident and early-career FPs included in our analysis.

**Table 1.** Planned practice characteristics of resident FP participants choosing to focus their practice (N=22).

| Characteristic |  | N(%) |
| --- | --- | --- |
| **Province** |  |  |
| British Columbia |  | 7 (32) |
| Ontario |  | 6 (27) |
| Nova Scotia |  | 9 (41) |
| **Planned area of clinical practice\*** |  |  |
| Focused practice |  | 22 (100) |
|  | Emergency medicine | 13 (59) |
|  | Hospitalist medicine | 9 (41) |
|  | Addiction medicine | 7 (32) |
|  | Sexual and reproductive health | 4 (18) |
|  | Obstetrics and maternity care | 3 (14) |
|  | Medical aesthetics, dermatology | 3 (14) |
|  | Geriatrics | 2 (9) |
|  | Mental health | 2 (9) |
|  | Palliative care | 2 (9) |
|  | Sports medicine | 2 (9) |
|  | Oncology | 1 (5) |
| Comprehensive family medicine practice |  | 21 (95) |
|  | Comprehensive family medicine practice | 13 (59) |
|  | Comprehensive family medicine practice with no obstetrics/prenatal care | 5 (23) |
|  | Comprehensive family medicine practice for a particular population | 3 (14) |

\*Planned areas of clinical practice are not mutually exclusive as most participants' intended practices incorporated a combination of these elements

**Table 2.** Practice characteristics of early-career FP participants with focused practices (N=38).

| Characteristic |  | N (%) |
| --- | --- | --- |
| Province |  |  |
| British Columbia |  | 14 (37) |
| Ontario |  | 13 (34) |
| Nova Scotia |  | 11 (29) |
| Area of clinical practice* |  |  |
| Focused practice |  | 38 (100) |
|  | Hospitalist medicine | 14 (37) |
|  | Emergency medicine | 12 (32) |
|  | Sexual and reproductive health | 9 (24) |
|  | Obstetrics and maternity care | 5 (13) |
|  | Surgical/procedural medicine | 4 (11) |
|  | Addiction medicine | 3 (8) |
|  | Medical Assistance in Dying (MAiD) | 3 (8) |
|  | Travel medicine | 2 (5) |
|  | Urgent care | 2 (5) |
|  | Student health, youth mental health | 2 (5) |
|  | Consultation for provincial workers' safety board | 1 (3) |
|  | Medical aesthetics, dermatology | 1 (3) |
|  | Palliative care | 1 (3) |
|  | Sports medicine | 1 (3) |
| Comprehensive family medicine practice |  | 30 (79) |
|  | Comprehensive family medicine practice | 22 (58) |
|  | Comprehensive family medicine practice with no obstetrics/prenatal care | 4 (11) |
|  | Comprehensive family medicine practice for a particular population | 2 (5) |
| Non-clinical |  | 2 (2) |
|  | Academic/administrative | 2 (2) |

\*Areas of clinical practice are not mutually exclusive as most participants' practices incorporated a combination of these elements

Among the resident FPs who intended to focus their practices, 21 (95%) anticipated practicing some form of comprehensive family medicine with focused practice. One resident FP envisioned solely working in focused practice. Among early-career FPs, 21 (55%) devoted more than half or all their time to a focused area and 30 (79%) offered some form of comprehensive family medicine. The most commonly reported areas of focused areas of practice in both groups were emergency and hospitalist medicine. Focused practice choices occurred on a continuum, ranging from the provision of all services under the umbrella of a defined area (e.g., dermatology) to a specific procedure within a particular area (e.g., only Botox injections).

### Key factors contributing to intentions or choices of focused practice

We identified three key and three minor themes of influential factors that helped explain participants’ decisions to pursue focused practice. Key themes were prominent across both resident and early-career FP datasets, while minor themes were less salient in the data. Themes are described in detail in Table 3 with illustrative quotations.

**Table 3.** Themes of influential factors for focused practice

| Themes | Group | Influence on choices of focused practice | Illustrative quotes |
| --- | --- | --- | --- |
| **Self-preservation within the current health care system** | *Resident FPs* | Plans to pursue focused practice due to: feeling undervalued and uncertain about future provincial policy changes; seeing the health care system as inadequately supporting and compensating comprehensive family medicine practice for the long hours and heavy workload, particularly with respect to the care of older and increasingly complex patients; significant overhead costs in comprehensive family medicine practice; and feeling that comprehensive family medicine practice involves detrimental impacts on their personal well-being (e.g., burnout) and work-life balance. | “It’s a bit of a crisis. I feel like a lot of physicians are burnt out ... And, you know, documentation also takes up time with forms and everything. And I feel like... that’s not really being considered. And when it comes to the fee-for-service model, that’s why I don’t think it would work for me just because patients are a bit more complex than they used to be. They’re living longer. There’s more drugs out there. So I’d like to kind of see like a nice balance. Like I don’t think you should be rushing through your patients or just having single issue appointments ... So I think when they’re [the government] making their policies and doing the compensation and payment plans, I’d like to see them sort of consider that...” R14, NS<br>“It basically comes back to what I was saying earlier with overhead. Overhead is not something that you have when you work as a hospitalist .... So you definitely make more money than you would in a clinic setting in a big city... the main thing about hospitalist work is that again it doesn’t come attached with you taking care of an office, of a staff. You have no overhead ... And then once you’re done your week of work, you don’t have the patients to follow after that.” R2, BC<br>“You’re always responsible for all the blood work coming in, all the x-rays that are done, new cancer diagnoses that are seen on CT scans. So having a clinic, it’s not like an 8 to 4 job. It’s a 24 hour job ... And then if you want to take vacation, you need someone to cover your patients while you’re gone ... I have a life and I have lots of interests outside of medicine. And so that impacts the career that I choose. Because I don’t want to go into a family medicine job where I have to work 80 hours a week and can’t do anything else.” R13, NS |
|  | *Early-career FPs* | Choosing to focus practice due to: perceptions of the government’s primary care policies undervaluing family physicians and health care quality, and inadequately compensating comprehensive family medicine for the long hours and heavy workload; beliefs that policy reform is necessary to increase the feasibility of comprehensive family medicine practice given the aging | “There’s such a huge generational gap in medicine. And you know, the generation that by and large is training us just doesn’t see another way to be .... But they truly think ... that people doing focused practices are providing inferior care ... I think it’s the entire sort of paradigm of the leadership generation in medicine right now is holding onto something that doesn’t exist anymore ... This generation of doctors, we’re not lazy and we don’t not care about patients. We’re just not willing to ruin the rest of our lives for the career. And we don’t think that that’s a reasonable expectation of us. Because, you know, you see it all the time and you hear it all the time. There’s always a letter to the editor about the young doctors not doing this, and the young doctors not doing that. And it’s self-preservation. We care about people too. We [are] also not willing to lay down our lives for the system.” FP4, BC |
|  |  | population and increasing complexity of disease; significant overhead costs in comprehensive family medicine practice; feeling pressure to work in an unsustainable, antiquated family physician role; and perceptions of comprehensive family medicine practice having detrimental impacts on participants' personal well-being and work-life balance. | "From what I see, there's a shortage of family doctors who want to work in family medicine. And I think compensation is part of it. But when I think about how I could do something like that for myself, I actually don't see the financial benefit in opening up my own practice unless I'm able to employ plenty of doctors ... It is not a risk I'm willing to take right now... I know that I feel like I'm not being compensated fairly to an extent. ... I just feel like for the work that we do, and the amount of responsibility and the amount of liability, and the amount of training we've gone through, I just don't feel like it's enough. And I don't see the Ministry ... doing enough especially for non-specialist physicians." FP18, ON<br>"If I were to take something like maternity leave...that does affect where you don't want to have 2,000 patients [in a comprehensive family medicine practice] and then have to go off for a year ... or however long you're on maternity leave. And so that would make me ... kind of think like do I actually want to take on patients? Or is that something I'd want to do after, you know, in 10 years when I feel like I've had a family and I'm back to working full-time? Or is it something that I just don't want to do because as soon as you have a roster of patients, it makes it very difficult to leave or to move or to change your mind as much... Like I would like to have more flexibility in terms of taking time off ... And finding locums is a little challenging..." FP26, BC |
| **Access to a support system** | *Resident FPs* | Intentions to pursue focused practice due to perceptions that focused areas often have an interdisciplinary team environment where collective expertise enables improved patient care, collegiality, and positivity in the work environment. | "Yeah, so the areas I'm interested in, I'm very much interested in women's health. And I do that now and will continue to do that ... I think just everybody there. They're in a good headspace. There's a low level of burnout and a high level of awareness around burnout. So people are good at supporting each other. People are kind." R3, BC<br>"Working in hospitals, I think it's a huge advantage over working in clinics in terms of multidisciplinary work. You know, in hospital, you basically have all the different specialties, right. You're going to have OT, you're going to have PT, dietician, SLP – everyone's around you. Which is awesome and I kind of like that teamwork. Whereas clinics, when you work in a family practice office, unless it's a big clinic and they have the multidisciplinary team, I find most clinics will have maybe one nurse or two. ... So yes, of course, I really like working with other specialists. I think it makes your life much easier and it helps us to provide better care. And it's one of the reasons why hospitalist, for example, is more attractive to me." R2, BC |
|  | *Early-career FPs* | Decisions to focus practice due to: role modelling from peers demonstrating the possibilities and benefits of focused practice; cross-coverage through call groups; and perceptions that focused areas often | "But I also have a really awesome [obstetrics] call group. So that on weekends, if I want to take the weekend off and have a weekend with my family, and not have to worry about being pulled away, that I can sign out to them. And I have a group of physicians that will care for my patients. I trust that my patients are in really good hands. ." FP10, NS |
|  |  | have a team environment where collective expertise enables improved patient care, collegiality, and decreased isolation. | “I actually enjoy being a hospitalist a great deal. I like family medicine in clinic but I really, really enjoy inpatient medicine... It’s a bit of a different collegiality because you’re working with people more directly. Whereas here [in the clinic], we each see our patients. And if I need to ask someone to come help me, they’ll come help me. But realistically they’re busy seeing their own patients.” FP33, ON |
| **Training experiences** | *Resident FPs* | Plans to focus practice due to: clinical training experiences revealing areas of interest; role modelling from mentors demonstrating the possibilities and benefits of focused practice; and encouragement from mentors to explore clinical interests. | “My residency program really got me passionate about helping people who live in the inner city areas, and people who have addiction issues. And then the other thing is transgender care. ... I didn’t even really know much about it. And it wasn’t really on my radar at all. So I was exposed to that through my [community health centre] rotation. And I just thought it was just a wonderful experience. So yeah, I continued to do some of that work throughout the year. And that’s why I would want to keep doing it.” R7, BC |
|  | *Early-career FPs* | Choosing to focus practice due to: clinical training experiences revealing areas of interest; role modelling from mentors demonstrating the possibilities and benefits of focused practice; and throughout training, developing perceptions of comprehensive family medicine practices as unappealing. | “It felt most of the doctors that I followed [in medical school], you know, would see 30 to 40 patients a day. They were mostly older, white men ... the physician I followed would see up to 50 patients a day... And it was exhausting. And I don’t think I saw myself in a model like that. And so even though I chose family, I think in my mind I knew I wasn’t going to practice in that manner. And I thought, well, maybe that means I’m just going to focus my practice in smaller areas, such as women’s health or join a community centre or something along those lines.” F19, ON |
| **Alignment with skills or values, and professional satisfaction** | *Resident FPs* | Intentions for focused practice due to its alignment with participants’ skills or values; allowing for maintenance of skills in a specific area; and enabling professional satisfaction through filling a perceived gap in care, having variety in work environments, or being intellectually stimulated. | “... being able to go between multiple environments, it just appeals to kind of my personality. Also just having kind of a wide range of skills that I can continue to use. Like emergency medicine skills, inpatient skills with an outpatient population, that just appeals to kind of what my own personal interests are ... But there’s a lot of us in medicine who have... again, we have this kind of like career ADHD. We just can’t stay in the same place.” R18, ON |
|  | *Early-career FPs* |  | “[What is most important to me in my career is] That I feel good about the work that I’m doing. That I feel like I’m contributing to my community in a way that helps people ... Maybe just what was needed in our community. ... MAiD was something that was under-served. And the woman that was only doing it at the time was quite stressed out. And when she approached me, it just made sense to do it. So I think it’s what my community needs definitely influences what I would change my practice to.” F22, BC |
| **Personal lived experience** | *Resident FPs* | Intentions or choices of pursuing focused practice influenced by prior life experiences. | “As a teenager, late teenager, a lot of my friends got quite heavily into drugs and then selling and doing drugs ... And I think that’s a big reason why I’m drawn to addictions as well – watching them go through that and be arrested and go to jail. These people I’ve known for 7, 8, 9 years. And their life took a huge nosedive that they’re only now recovering from, is a big reason why I’m drawn to addictions.” R16, NS |
|  | *Early-career FPs* |  | “When I was in high school and I guess in undergrad as well, I did a lot of work at a nursing home. I did volunteer work at a nursing home. And I felt that that really actually had probably influenced my decisions to work as a hospitalist in family medicine, because I was always so exposed to the age group which I'm working with now. That's why I feel quite comfortable working in like a hospital or a nursing home setting. So those kinds of things in terms of a team-based setting and as well as being outside of the clinic. ... And I keep doing it to this day." FP38, ON |
| **Presence of existing opportunities in focused practice** | *Resident FPs* | Plans to focus practice due to feeling encouraged to practice in a focused area based on availability of work, community needs, or a high demand for FPs in a particular focused area due to limited specialist availability. | "For more specific things like dermatology, I know there's always sort of a very long wait list to see a dermatologist. So I think, at least from what I've seen, anyone who's kind of had a focused interest in that, there's no shortage of people or patients coming to see you." R8, NS<br>"The only thing like really that I thought I would consider is addictions. I think they're always hiring. And I don't know, maybe I'm a little naïve and I just have this sense that things are going to work out... But I think at least for the addictions part, I think there's enough jobs." R20, ON |
|  | *Early-career FPs* | Choosing to focus practice due to having many job opportunities within a particular focused practice area. | "Certainly there is no shortage of things being offered or opportunities. I think it's unlikely that it's not going to be a viable approach to practice [as a comprehensive family medicine locum and hospitalist]." FP12, NS<br>"And then I got married and we decided to pack up and move to [city] for a couple of years and, you know, just go...because my husband has a flexible job. And I figured if there's anywhere else I could work full-time in sexual health, it's probably [city]." FP4, BC |

#### Self-preservation within the current structure of the health care system

Both participant groups described issues within the health care system that influenced their choices of focused practice, specifically with regards to remuneration and workload. Certain physician remuneration models, such as fee-for-service, deterred participants from practicing comprehensive family medicine. Fee-for-service was seen as inadequately compensating for the long hours, workload, and overhead costs associated with longitudinal care for increasingly complex patients. In contrast, focused practice was seen as more attractive and sustainable due to better compensation and fewer administrative costs.

The early-career FPs in our study described policies governing primary care delivery in all three provinces as contributing to heavy workloads and concerns about burnout. Similarly, resident FPs relayed observations of FP mentors being overworked, inadequately remunerated, and having difficulty securing time off in comprehensive family medicine practice. In contrast, both participant groups felt that focused practice offered better remuneration and flexibility to choose hours worked, allowing more time for family commitments, hobbies, or parental leave. Resident and early-career FPs saw parental leave as incompatible with comprehensive family medicine practice due to challenges securing locum coverage, perceived resentment from patients for time off, and interruptions in patient continuity of care.

Further, early-career FPs with focused practices described feeling pressured during their training to work in what they considered an antiquated FP role. They shared that instructors put emphasis on a paradigm of comprehensive family medicine practice that involved working around the clock to serve patients and that this was the best way to practice. Early-career FP participants perceived these traditional comprehensive FP roles as unachievable for current and future levels of patient complexity and need, and detrimental to their well-being and family. Resident and early-career FPs alike expressed an unwillingness to sacrifice work-life balance, believing that policy reform was necessary for them to consider a broader scope of practice. Both participant groups were unanimously dissatisfied with provincial government policies, and considered their governments to be unresponsive to their needs and undervaluing FPs.

#### Access to a support system

Resident and early-career FPs felt focused practice offered greater access to a support system compared to comprehensive family medicine practice. Both participant groups viewed call groups and team-based care environments within focused practice areas (e.g., hospitalist medicine) as support systems that improved quality of care, facilitated knowledge sharing, and decreased isolation. Such systems facilitated self-preservation in the current health care system. Early-career FPs also described their peers as role models who demonstrated the feasibility of incorporating focused areas into their overall practices.

#### Training experiences

Both participant groups reported training experiences that increased their comfort with focused areas of practice and created recognition workload in comprehensive family medicine was not an ideal match for their desired lifestyle. This belief was reinforced by resident and early-career FP perceptions that their mentors were exhausted in comprehensive family medicine practice environments.

### Minor themes for influential factors for focused practice

Resident and early-career FPs described feeling attracted to a particular focused area (e.g., hospitalist medicine) because it aligned with their skills/values or helped them maintain specific competencies. Both participant groups indicated that a focused practice brought with it a sense of professional satisfaction by filling a perceived gap in care, increasing variety in their work, or being intellectually stimulated.

Both participant groups also described personal lived experiences that sparked their interest in particular focused practice areas. For example, experiences with family members, friends, or community members with mental health struggles or addictions, along with prior volunteer experiences, contributed to interest in focused practices in mental health and addictions.

Resident FPs described being attracted to focusing their practice due to available opportunities, community needs, and limited specialist availability. Similarly, early-career FPs reported being inclined to incorporate focused areas into their practices due to a multitude of job opportunities in focused practice.

### Comparison between provinces

Our results were similar across the provinces studied. Resident and early-career FPs in British Columbia and Nova Scotia described similar concerns about inadequate compensation for the workload and responsibility involved in comprehensive family medicine practice. Specifically, early-career FPs in these provinces desired fee-for-service fee schedules that aligned with other provinces or alternative payment models. In Ontario, resident and early-career FPs expressed dissatisfaction with the province’s numerous payment models, describing loss of control over earnings in comprehensive family medicine practice, uncertainty, and distrust due to a fluctuating policy landscape (e.g., fee cuts, role restrictions).

## Interpretation

The resident and early-career FPs with focused practices found focused practice attractive for numerous reasons, including: more manageable workloads, better remuneration, and improved work-life balance; familiarity from prior exposure during training; and the presence of a supportive team environment. Less common reasons for opting for focused practice included alignment with participants’ skills, values, or an ability to feel professional satisfaction; personal lived experience; and having a multitude of opportunities to practice. Resident and early-career FPs described focused practice as a way to circumvent burnout or exhaustion, which they considered to be an untenable component of comprehensive family medicine practice in the current health care system. Discontent with provincial policies and the lack of government responsiveness to their concerns was apparent across all provinces and practice types.

### Explanation of the findings

Previous work has identified newly graduating resident FPs as more likely to intend to provide a broad scope of practice compared to FPs in current practice.[3] Though our study was not designed for statistical comparisons, we also found that resident FPs were more likely to report intending to practice comprehensive family medicine than early-career FPs. The post-training working environment and lack of support for providing a broad scope of services have been suggested as possible driving forces for this finding.[3] Other studies have described comprehensive family medicine practice as too broad, overwhelming, involving a high degree of responsibility, allowing for minimal work-life balance,[7, 8] and providing insufficient financial incentives.[9] Factors reported to support the choice of focused practice include superior financial incentives,[14] opportunities for intellectual stimulation,[7, 8] reduced stress from a lower workload,[7] and prior training exposures.[24] Our study confirms these findings. Canadian FPs have reported feeling unprepared to deliver comprehensive care.[12, 25] We did not find this in our study. Instead, we found system-level barriers, linked to government policy, influencing focused practice choices.

Our study confirms influential factors for focused practice previously identified in the literature, and describes additional elements that, to our knowledge, have not been described before. Moreover, we found similar factors contributing to choices of focused practice for both resident and early-career FPs, demonstrating that these factors are consistent prior to and during independent practice.

### Future directions

There is contention about the benefits and harms of increasing focused practice within family medicine.[26] The impact of rising numbers of FPs practicing in focused areas on the supply of FPs providing comprehensive care is still unknown and requires more research. Further work is also needed to identify reforms that may encourage FPs to offer a comprehensive scope of family medicine while supporting their personal and professional well-being.

### Limitations

Data collection for this study occurred prior to the onset of COVID-19 and therefore does not necessarily reflect the current environment. This study only includes individuals who responded to requests to participate, which may not reflect all types of resident and early-career FPs. We did not ask participants specific questions about sex/gender, geography, and training location, focusing instead on open-ended questions. We may have developed a richer description by probing these additional areas.

## Conclusions

Numerous elements, including system-level issues, shape resident and early-career FP choices to narrow the scope of their practices. Our work found similar influential factors shaping practice choices among both resident and early-career FPs in three Canadian provinces. Further work is needed to understand the potential impact of focused practice on the delivery of health care services in Canada, and associated policy implications.

## Supporting information

COREQ checklist

## Data Availability

Due to the identifiable nature of raw data (interview transcripts), they are not publicly available. Requests for additional information about study data or results can be forwarded to the corresponding author (AG).

## Appendices

Appendix 1. Screening survey.

Appendix 2. Interview guide for resident and early-career family physicians.

## Ethical approval

This study was approved by the Simon Fraser University (#H18-03291), University of Ottawa (#S-05-18-776), and Nova Scotia Health Authority research ethics boards (#1023561).

## References

1. Chan BTB. The declining comprehensiveness of primary care. CMAJ. 2002;166(4):429–34.

2. Collier R. The changing face of family medicine. CMAJ. 2011;183(18):E1287–E8.

3. Coutinho AJ, Cochrane A, Stelter K, Phillips RL, Peterson LE. Comparison of intended scope of practice for family medicine residents with reported scope of practice among practicing family physicians. JAMA. 2015;314(22):2364.

4. Freeman TR, Boisvert L, Wong E, Wetmore S, Maddocks H. Comprehensive practice: Normative definition across 3 generations of alumni from a single family practice program, 1985 to 2012. Can Fam Physician. 2018;64(10):750–9.

5. Reitz R, Horst K, Davenport M, Klemmetsen S, Clark M. Factors Influencing Family Physician Scope of Practice: A Grounded Theory Study. Fam Med. 2018;50(4):269–74.

6. Schultz SE, Glazier RH. Identification of physicians providing comprehensive primary care in Ontario: a retrospective analysis using linked administrative data. CMAJ Open. 2017;5(4):E856–E63.

7. Beaulieu M-D, Rioux M, Rocher G, Samson L, Boucher L. Family practice: Professional identity in transition. A case study of family medicine in Canada. Soc Sci Med. 2008;67(7):1153–63.

8. Beaulieu MD, Dory V, Pestiaux D, Pouchain D, Rioux M, Rocher G, et al. What does it mean to be a family physician?: Exploratory study with family medicine residents from 3 countries. Can Fam Physician. 2009;55(8):e14–20.

9. Calnan M. Variations in the range of services provided by general practitioners. Fam Pract. 1988;5(2):94–104.

10. Dhillon P. Shifting into third gear: current options and controversies in third-year postgraduate family medicine programs in Canada. Can Fam Physician. 2013;59(9):e406–12.

11. Lerner J. Wanting family medicine without primary care. Can Fam Physician. 2018;64(2):155–6.

12. Vogel L. Are enhanced skills programs undermining family medicine? CMAJ. 2019;191(2):E57–E8.

13. Grierson L, Vanstone M, Alice I. Understanding the Impact of the CFPC certificates of Added Competence [Internet]. 2016 [cited 2021 April 28]. Available from: https://www.cfpc.ca/CFPC/media/PDF/2020-04-CAC-Impact-Study-Report.pdf.

14. Glazer J. Specialization in family medicine education: abandoning our generalist roots. Fam Pract Manag. 2007;14(2):13–5.

15. Lavergne MR, Goldsmith LJ, Grudniewicz A, Rudoler D, Marshall EG, Ahuja M, et al. Practice patterns among early-career primary care (ECPC) physicians and workforce planning implications: protocol for a mixed methods study. BMJ Open. 2019;9(9):e030477.

16. Starfield B. Primary Care: Concept, Evaluation, and Policy. New York: Oxford University Press; 1992.

17. Institute of Medicine. Primary care: America’s health in a new era. Washington, D.C.: National Academy Press; 1996.

18. Braun V, Clarke V. Using thematic analysis in psychology. Qual Res Psychol. 2006;3(2):77–101.

19. Aronson J. A pragmatic view of thematic analysis. The Qualitative Report. 1993;2(1):1–3.

20. QSR International Pty Ltd. NVivo 12 [Software]. 2018 [cited 2021 March 19]. Available from: https://www.qsrinternational.com/nvivo-qualitative-data-analysis-software/home.

21. Guest G, MacQueen K, Namey E. Comparing Thematic Data. 2012. In: Applied Thematic Analysis [Internet]. Thousand Oaks, California: SAGE Publications, Inc.; [161-86]. Available from: https://methods.sagepub.com/book/applied-thematic-analysis.

22. Creswell J, Poth C. Standards of validation and evaluation. Qualitative inquiry and research design: Choosing among five approaches. Thousand Oaks, CA: Sage Publications; 2018. p. 253–86.

23. Carter N, Bryant-Lukosius D, DiCenso A, Blythe J, Neville AJ. The use of triangulation in qualitative research. Oncol Nurs Forum. 2014;41(5):545.

24. Coutinho AJ, Levin Z, Petterson S, Phillips RL, Peterson LE. Residency program characteristics and individual physician practice characteristics associated with family physician scope of practice. Academic medicine : journal of the Association of American Medical Colleges. 2019;94(10):1561–6.

25. Osborn R, Moulds D, Schneider EC, Doty MM, Squires D, Sarnak DO. Primary care physicians in ten countries report challenges caring for patients with complex health needs. Health Aff. 2015;34(12):2104–12.

26. Collier R. A comprehensive view of focused practices. CMAJ. 2011;183(18):E1289–90.

