## Appendices for "Factors influencing focused practice: A qualitative study of resident and early-career family physician practice choices"

#### Appendix 1: Screening Surveys for family physicians and residents

##### Resident Family Physician Survey

1) What is your gender?
   - ☐ Female
   - ☐ Male
   - ☐ Non-binary / third gender
   - ☐ Prefer to self-describe
   - ☐ Prefer not to answer

2) What is your current relationship status?
   - ☐ Single/Divorced/Separated/Widowed
   - ☐ Married/Common-law/Life Partner
   - ☐ Other, please specify: _______________
   - ☐ Prefer not to answer

3) Do you have any dependents (e.g., child, parent, grandparent)?
   - ☐ Yes – child(ren)
   - ☐ Yes – adult(s)
   - ☐ Yes – both
   - ☐ No

4) Is there another person (non-dependant adult) in your life who influences decisions you make about how and where you practice?

   ☐ Yes ☐ No

5) Where did you graduate from medical school?
   - ☐ Canadian Medical Graduate
   - ☐ International Medical Graduate

6) Which of the following best describe the community(ies) in which you are doing your residency? Please select all that apply.

- ☐ Inner city
- ☐ Urban/suburban
- ☐ Small town
- ☐ Rural
- ☐ Remote
- ☐ Mixture of environments

7) Which of the following best describes the organizational model(s) in which you practice? Please select all that apply.

- ☐ Solo practice
- ☐ Group physician practice
- ☐ Interprofessional team-based practice
- ☐ Other, please specify: _________________

8) Which best describes the kind of practice you intend to have after your residency?

- ☐ Comprehensive practice
- ☐ Special interest practice (less than 50% of the time on specialized care)
- ☐ Focused practice (50% or more of the time on specialized care)
- ☐ Other, please specify: _________________

### Early-Career Family Physician Survey

1) What is your gender?
   - ☐ Female
   - ☐ Male
   - ☐ Non-binary / third gender
   - ☐ Prefer to self-describe
   - ☐ Prefer not to answer

2) What is your current relationship status?
   - ☐ Single/Divorced/Separated/Widowed
   - ☐ Married/Common-law/Life Partner
   - ☐ Other, please specify: _______________
   - ☐ Prefer not to answer

3) Do you have any dependents (e.g., child, parent, grandparent)?
   - ☐ Yes – child(ren)
   - ☐ Yes – adult(s)
   - ☐ Yes – both
   - ☐ No

4) Is there another person (non-dependant adult) in your life who influences decisions you make about how and where you practice?

   ☐ Yes ☐ No

5) Where did you graduate from medical school?
   - ☐ Canada
   - ☐ Outside of Canada

6) Which of the following best describe the community(ies) in which you predominantly practice? Please select all that apply.
   - ☐ Inner city
   - ☐ Urban/suburban
   - ☐ Small town
   - ☐ Rural
   - ☐ Remote
   - ☐ Mixture of environments

7) Which of the following best describes the organizational model(s) in which you practice? Please select all that apply.

- ☐ Solo practice
- ☐ Group physician practice
- ☐ Interprofessional team-based practice
- ☐ Other, please specify: _________________

8) Which of the following best describes your practice?

- ☐ Comprehensive practice
- ☐ Special interest practice (less than 50% of the time on specialized care)
- ☐ Focused practice (50% or more of the time on specialized care)
- ☐ Other, please specify: _________________

9) Which of the following are methods by which you receive payment? Please select all that apply.

- ☐ Fee-for-service
- ☐ Salary
- ☐ Capitation
- ☐ Sessional/per diem/hourly
- ☐ Service contract
- ☐ Blended
- ☐ Other, please specify: _________________

### Appendix 2: Interview Guides for Resident and Early-Career Family Physicians

#### Interview Guide: Resident Family Physicians

1. Can you tell me a little bit about where you're at in your residency right now?
   - *Prompt*: Which year of residency are you in?

##### Future Practice Description

2. How do you see your practice unfolding when you finish residency?

3. Long-term, what would you like your practice to look like?
   - Where would you like to practice?
   - How would the practice be organized?
     - *Prompt*: practice models (solo, group, interprofessional) and types (comprehensive, special interest, focused)
   - What types of clinicians or other allied health professionals would you like to work with and how?
     - *Prompt*: primary care providers including other types of physicians, nurses, pharmacists, maybe social workers
   - Do you have a preferred compensation type?
     - *Prompt*: fee-for-service, salary, capitation, sessional/hourly, service contract, blended
   - What would you like your work week to look like?

4. Tell me about any particular clinical interests you have as a family physician.
   - Which patient populations are you interested in?
   - How do you hope to incorporate these interests into your practice?
   - Do you see this changing over time?

5. Have you had any experience in a practice like the one you described? What did you like and not like about it.

6. How do you see your practice changing over time?
   - If IMG with a return of service: how will your practice change after your return of service is satisfied?

   [If participant mentions "comprehensive" probe: what does comprehensive mean for you?]

##### Priorities

7. When you think about your career, what is most important to you?

8. In what ways, if any, do your personal priorities or goals influence your career plans?
   - How might your personal relationships influence your career plans?
   - How might parenthood or caregiving influence your career plans?
   - How might financial considerations influence your career plans?
   - How might your gender influence your career plans?

- How might your other personal characteristics influence your career plans?

9. Are your career plans influenced by anything other than your personal priorities and goals?

#### Past experiences

10. How did your medical school experience influence your career plans as a family physician? (*Re-direct away from responses about why family medicine was chosen as a specialty)*
    - Positive or negative experiences?
    - Did you have any experiences during medical school that stand out?
    - Did any key people influence your plans?

11. How did your residency influence your career plans as a family physician?
    - Positive or negative experiences?
    - Did you have any experiences during your residency that stand out?
    - Did any key people influence your plans?

12. Tell me about your CaRMs experience.
    - Was family medicine your first choice?
    - Did you have to make trade-offs between specialty and location of residency?

13. Tell me about any other life experiences you've had that influence your career plans as a family physician.

#### Practice Opportunities

14. At the start of the interview, you told me what you would like your clinical practice to look like. Do you expect you will be able to achieve this ideal type of practice? Why/Why not?
    - Are opportunities available for this type of practice?
    - Are there restrictions or barriers to you having this ideal practice?
    - How will you populate your practice?
    - How do your gender or other personal characteristics impact your ability to achieve this type of practice?
    - *(Prompt if needed)* Are you planning to stay in the same province where you are completing your residency?

*Ontario specific question*

15. In what ways, if any, do you anticipate provincial policies such as managed entry into team-based practices will affect your practice choices?

#### Wrap up

16. If you were mentoring a new family medicine resident, what advice would you give them about planning their career in family medicine?

17. Anything else that you think is important for me to know?

### Interview Guide: Early-Career Family Physicians

#### Current & Future Practice Description

1. How did your career unfold out of residency?

2. Tell me briefly about your current practice.
   - How is your practice organized?
   - Who else do you work with?
   - How are you (and your team) compensated?
   - What does your work week look like?

3. Tell me about any particular clinical interests that you have as a family physician.
   - Which patient populations are you interested in?
   - How have you incorporated these interests into your practice?
   - Do you see this changing over time?

4. In what ways, if any, does your current practice differ from your ideal type of practice?

5. How do you see your practice changing over time?
   - If IMG with a return of service: how will your practice change after your return of service is satisfied?

6. [If participant mentions "comprehensive" probe: what does comprehensive mean for you?]

#### Priorities

7. When you think about your career, what is most important to you?

8. In what ways, if any, did your personal priorities or goals influence your career?
   - How did your personal relationships influence your career?
   - How did parenthood or caregiving influence your career?
   - How did financial considerations influence your career?
   - How did your gender influence your career?
   - How did your other personal characteristics influence your career plans?

9. (if no exogenous factors emerge in Q 4, 5, 6) What kinds of other influences have you experienced or anticipate that may influence your practice changes over time?
   i. E.g. Community, professional, regulatory influences

#### Past Experiences

10. How did your medical school experience influence your career plans as a family physician? (*Re-direct away from responses about why family medicine was chosen as a specialty)*
    - Positive or negative experiences?
    - Did you have any experiences stand out in primary care during training?
    - Did any key people influence your plans?

11. How did your residency influence your plans for family practice?
    - Positive or negative experiences?
    - Did you have any experiences in primary care stand out during training?
    - Did any key people influence your plans?

12. Tell me about your CaRMs experience
    - Was family medicine your first choice?
    - Did you have to make trade-offs between specialty and location of residency?

13. Tell me about any other life experiences you've had that influence your career plans as a family physician?

#### Why: Policy Environment and Practice Opportunities

14. At the start of the interview, you told me about how you practice now and what you would like your clinical practice to look like. Do you expect you will be able to achieve this ideal type of practice? Why / why not?
    - Are opportunities available for your preferred type of practice?
    - Are there restrictions or barriers to you having this ideal practice?
    - How will you populate your practice?
    - How do your gender or other personal characteristics impact your ability to achieve this type of practice?

#### Wrap up

15. If you were mentoring a new family medicine resident, what advice would you give them about planning their career in family medicine?

16. Anything else that you think is important for me to know?
