## Supplementary material for "Factors influencing focused practice: A qualitative study of resident and early-career family physician practice choices": COREQ checklist

### COREQ Checklist – dated May 14, 2021

| **Item** | **Guide questions/description** | **Response and page number referenced** |
|---|---|---|
| *Domain 1: Research team and reflexivity* | | |
| 1. Interviewer/ facilitator | Which author/s conducted the interview or focus group? | Page 6:<br>"One research analyst (ER, LJ, MM) per province conducted one-on-one semi-structured, in-depth interviews." |
| 2. Credentials | What were the researcher's credentials? e.g. PhD, MD | Research credentials are provided in the author list (page 1). |
| 3. Occupation | What was their occupation at the time of the study? | Researchers completing data collection and analysis were employed as research analysts for the study. |
| 4. Gender | Was the researcher male or female? | All three research analysts were female. This detail was not included in the manuscript as the interviewer's gender did not have a bearing on the content of interviews. |
| 5. Experience and training | What experience or training did the researcher have? | Page 6:<br>"Each research analyst was trained in qualitative interviewing." |
| 6. Relationship established | Was a relationship established prior to study commencement? | N/A |
| 7. Participant knowledge of the interviewer | What did the participants know about the researcher? e.g. personal goals, reasons for doing the research | Page 5:<br>"Participants were provided with information about the study, the study rationale, and the names of research team members." |
| 8. Interviewer characteristics | What characteristics were reported about the interviewer/facilitator? e.g. Bias, assumptions, reasons and interests in the research topic | Page 5:<br>"We provided participants with study information and an honorarium."<br><br>Study information included a list of the research team members. |
| *Domain 2: Study design* | | |
| 9. Methodological orientation and Theory | What methodological orientation was stated to underpin the study? e.g. grounded theory, discourse analysis, ethnography, phenomenology, content analysis | Page 7:<br>"We used iterative, inductive thematic analysis.[18]"<br><br>Page 7:<br>"We conducted comparative analysis[21] to compare thematic patterns identified from the early-career FP transcripts with the resident FP dataset." |
| 10. Sampling | How were participants selected? e.g. purposive, convenience, consecutive, snowball | Page 5:<br>"We used purposeful sampling to maximize variation across self-identified gender, marital status, dependents, training location, years of training, years |

| | | |
|---|---|---|
| | | in practice, scope of practice, and practice models in each province.” |
| 11. Method of approach | How were participants approached? e.g. face-to-face, telephone, mail, email | Page 5:<br>“We recruited resident and early-career FPs for the qualitative arm of this larger mixed methods study through family medicine residency programs, social media, research conferences, and selected provincial medical associations.” |
| 12. Sample size | How many participants were in the study? | Page 5:<br>“We interviewed 31 of 32 resident FPs and 63 of 69 early-career FPs who had been invited to participate in the larger mixed methods study.”<br><br>Page 6:<br>“The sample used in this paper consists of 22 resident FPs and 38 early-career FPs who: i) self-identified in the screening survey as intending to practice or currently practicing within a focused area, and/or ii) described focused practice elements in their overall practice during the interview.” |
| 13. Non-participation | How many people refused to participate or dropped out? Reasons? | Page 5:<br>“Reasons for nonparticipation included scheduling conflicts (n=2), no response (n=4), or withdrawal with no reason provided (n=1).” |
| 14. Setting of data collection | Where was the data collected? e.g. home, clinic, workplace | Page 6:<br>“Telephone interviews were conducted using a semi-structured interview guide specific to each subgroup (see Appendix 2).”<br><br>Given that interviews were conducted via telephone or videoconference, participants were able to participate from a location of their choosing. |
| 15. Presence of non-participants | Was anyone else present besides the participants and researchers? | Page 6:<br>“One research analyst (ER, LJ, MM) per province conducted one-on-one semi-structured, in-depth interviews.” |
| 16. Description of sample | What are the important characteristics of the sample? e.g. demographic data, date | Planned practice characteristics of resident family physicians are presented in Table 1 (page 9).<br><br>Practice characteristics of early-career family physicians are presented in Table 2 (page 10). |

| | | |
|---|---|---|
| 17. Interview guide | Were questions, prompts, guides provided by the authors? Was it pilot tested? | Page 5:<br>“Telephone interviews were conducted using a semi-structured interview guide specific to each subgroup (see Appendix 2).” |
| 18. Repeat interviews | Were repeat interviews carried out? If yes, how many? | N/A |
| 19. Audio/visual recording | Did the research use audio or visual recording to collect the data? | Page 6:<br>“Interviews were audio-recorded and transcribed verbatim.” |
| 20. Field notes | Were field notes made during and/or after the interview or focus group? | Page 6:<br>“Research analysts recorded their reflections and interview summaries after each interview.” |
| 21. Duration | What was the duration of the interviews or focus group? | Page 5: “Selected individuals were then invited to participate in a 60-minute interview.” |
| 22. Data saturation | Was data saturation discussed? | Page 6: “Participant recruitment occurred iteratively until no new themes were identified in interviews.” |
| 23. Transcripts returned | Were transcripts returned to participants for comment and/or correction? | Page 7:<br>“We used accepted strategies for validating qualitative research.[22] The findings were: i) triangulated across a large sample size of participants with diverse experiences from three provinces and two types of participant groups;[23] and ii) presented to FP members of the research team (DS, IS, GM), who were not involved in the analysis process for this paper, to confirm data interpretation.” |
| *Domain 3: Analysis and findings* | | |
| 24. Number of data coders | How many data coders coded the data? | Page 7:<br>“For the qualitative arm of the larger mixed methods study, three research analysts (ER, LJ, MM) with experience in qualitative analysis generated initial resident and early-career FP codebooks through inductive coding of one resident and one early-career FP interview.[19]” |
| 25. Description of the coding tree | Did authors provide a description of the coding tree? | Page 7:<br>“For the qualitative arm of the larger mixed methods study, three research analysts (ER, LJ, MM) with experience in qualitative analysis generated initial resident and early-career FP codebooks through inductive coding of one resident and one early-career FP interview.[19] Codebooks were then refined through application to a subset of transcripts |

| | | |
|---|---|---|
| | | with guidance from the senior author (AG).” |
| 26. Derivation of themes | Were themes identified in advance or derived from the data? | Page 7:<br>“We used iterative, inductive thematic analysis.[18] For the qualitative arm of the larger mixed methods study, three research analysts (ER, LJ, MM) with experience in qualitative analysis generated initial resident and early-career FP codebooks through inductive coding of one resident and one early-career FP interview.[19]” |
| 27. Software | What software, if applicable, was used to manage the data? | Page 7:<br>“The research analysts used the final codebook to code transcripts from their respective provinces in NVivo 12[20].” |
| 28. Participant checking | Did participants provide feedback on the findings? | N/A |
| 29. Quotations presented | Were participant quotations presented to illustrate the themes / findings? Was each quotation identified? e.g. participant number | Page 11:<br>“Themes are described in detail in Table 3 with illustrative quotations.”<br><br>Quotations in Table 3 are identified by participant number. |
| 30. Data and findings consistent | Was there consistency between the data presented and the findings? | The findings were derived from the data using an inductive approach (see page 7), and therefore are consistent with the data presented. |
| 31. Clarity of major themes | Were major themes clearly presented in the findings? | Page 11: “We identified three key and three minor themes of influential factors that helped explain participants’ decisions to pursue focused practice. … Themes are described in detail in Table 3 with illustrative quotations.”<br><br>Themes are clearly presented both in the body of the paper, and in Table 3, where representative participant quotations are provided for each theme. |
| 32. Clarity of minor themes | Is there a description of diverse cases or discussion of minor themes? | Page 11:<br>“We identified three key and three minor themes of influential factors that helped explain participants’ decisions to pursue focused practice. Key themes were prominent across both resident and early-career FP datasets, while minor themes were less salient in the data.” |
